# A mixed methods study exploring food insecurity and diet quality in households accessing food clubs in England

**DOI:** 10.1101/2024.12.03.24318378

**Authors:** Nida Ziauddeen, Elizabeth Taylor, Nisreen A Alwan, Fran Richards, Barrie Margetts, Tim Lloyd, Marta Disegna, Naomi Mason, Ravita Taheem, Dianna Smith

**Affiliations:** School of Primary Care, Population Sciences and Medical Education, Faculty of Medicine, University of Southampton, Southampton, UK; NIHR Applied Research Collaboration Wessex, Southampton, UK; School of Geography and Environmental Science, University of Southampton, Southampton, UK; University Hospital Southampton NHS Foundation Trust, Southampton, UK; PPI contributor; Faculty of Medicine, University of Southampton, Southampton, UK; Bournemouth University Business School, Bournemouth, UK; Dipartimento di Tecnica e Gestione dei Sistemi Industriali, Università degli Studi di Padova, Italy; Public Health Dorset, Dorset, UK; Southampton City Council, Civic Centre, Southampton, UK; NIHR Southampton Biomedical Research Centre, University of Southampton and University Hospital Southampton NHS Foundation Trust, Southampton, UK

**Author notes:** These authors contributed equally to this work (Nida Ziauddeen, Elizabeth Taylor). Corresponding author: Dianna Smith, School of Geography and Environmental Science, University of Southampton, Southampton, UK.

**Keywords:** food insecurity, diet quality, wellbeing, food aid

## Abstract

**Background:** Food clubs are a higher-agency food aid intervention that charge a small fee for a set number of items. Some incorporate longer-term solutions such as budgeting support and cooking skills. These are in place in England to help address inadequate reliable access to affordable, nutritious food. We used a convergent parallel mixed methods design to describe the food insecurity households accessing food clubs experience and to assess diet quality and wellbeing at the start and after at least three months of using food clubs in the South of England.

**Methods:** Participants accessing food clubs in Wessex from March 31 to November 3, 2022 were recruited after providing informed consent. They completed a survey at recruitment that collected data on diet and health. Food security was assessed using the modified six-item US Department of Agriculture (USDA) food security survey module, and wellbeing using the short form Warwick-Edinburgh Mental Wellbeing Scale (WEMWBS). Follow-up surveys were conducted after participants used the clubs for at least three months. Participants were invited to take part in a semi-structured interview.

**Results:** Of the 90 participants recruited at baseline, 52% were aged 35–54 years, 74% were female, 81% were of White ethnicity, and 71% reported having at least one dependent child. Food security status was calculated in 69 participants who answered all six questions of the USDA module, with 42% reporting low and 43% very low food security. Among participants with follow-up (n=52), low food security was 41% and very low food security was 18% at follow-up.

Eleven participants were interviewed. Two themes explored impact and experiences of food club. Impact illustrated how participants consumed a more varied diet, experienced less financial pressure, and improved health, wellbeing and social interaction. Experiences of food clubs explored limitations of time and food range at clubs, developing a sense of community and overcoming stigma.

**Conclusion:** This study is the first in the UK to explore potential diet, food security and wellbeing impacts of food clubs. Ongoing impact evaluation will enable optimisation of interventions for the populations they serve, such as inviting other organisations/groups to attend/be available for members.

## Introduction

Household food insecurity is a longstanding social inequality in many high-income countries (1). Sometimes referred to as food poverty in the UK, the term describes households that experience nutritionally inadequate diets in the form of reduced portions, poorer diet quality, or skipped meals most often due to financial pressures (2,3).These elements of food insecurity are reflected in standard questions asked in surveys which capture this phenomenon, such as the US Department of Agriculture’s (USDA’s) measure (4). The UK government applied a brief definition of household food security where households are ‘…considered to have sufficient, varied food to facilitate an active and healthy lifestyle” in a 2024 report, noting that 10% of UK households were food insecure (5). Extended definitions of food security account accessing food without accessing charity and note the challenge of uncertainty of having enough food (6).

Food insecurity has gained increasing attention over the last few decades in the UK (7). Historically, systematic data collection on the incidence of food insecurity in UK households was minimal, with this lack of evidence masking the scale of the problem. More recently, regular surveys of households have helped to fill the evidence gap (e.g. the biannual Food and You 2 survey, the annual Family Resource Survey and a quarterly survey commissioned by the Food Foundation). These data enable researchers and governments in the UK to identify the prevalence and dynamics of household food insecurity over time, highlighting the demographic characteristics of affected households.

In light of the emerging evidence on food insecurity, the primary policy response has been to provide additional funds via welfare benefits and other means of support, particularly for households with children, such as Free School Meals, the Household Support Fund and the Holiday activities and food programmes. However, the income threshold for these government interventions is very low^1^ leaving many households requiring additional or alternative forms of assistance. Emergency intervention typically takes the form of food banks, where free parcels of food are given to a person or household referred by another organisation, school, or General Practitioner. Usually there is little choice in terms of the food provided in these parcels as they are predominantly made up from donated or surplus food from suppliers including supermarkets or larger organisations that collate and redistribute surplus food (FareShare in the UK). Initially, food banks provided only ambient (tinned, dry) foods but many more now have some refrigeration capacity and offer chilled foods, alongside fresh or frozen produce. Many groups are calling for a cash-first response to food insecurity, to provide national funds to people and allow them to buy what is needed (8). One example of this was the £20 per week uplift in Universal Credit payments during the Covid-19 pandemic. This payment may have supported a reduction in the prevalence of food insecurity during this time (5).

There are known barriers to accessing food banks such as the stigma of accepting help, or where children are involved, a perceived fear among parents that intervention may risk their children being taken into care/social services (9–11). Alternatives to referral-only food banks include food pantries/clubs, where people pay a small fee each week to choose from a range of heavily discounted food or household items; community fridges, where anyone can come and collect free food; or box schemes, where for a small/no fee households receive a parcel of food, perhaps around a recipe, although there is typically no choice in the food available. Among these alternatives, the food pantry or club represents a higher-agency food aid intervention, where people are given greater autonomy over the food they receive unlike in traditional food banks. This type of intervention is often referred to as a choice model, since it allows clients to choose food rather than receive a pre-packed parcel (12–15). Despite the growing popularity of this model of food intervention, their potential impacts in terms of improving food security, diet quality and wellbeing are yet to be studied in the UK. Overall, there is growing awareness that food aid is forming a larger part of the food environment for some households, meaning that it is part of the resources available influencing what people consume (5,16,17).

Food insecurity is a public health issue, with negative impacts on physical and mental health (18). The food consumed in less food secure households is often of lower nutritional quality (2,3) as less processed foods (especially fresh produce, meat, dairy) are more expensive (19). Parents reducing their portion size or skipping meals to ensure their children have more food is a common occurrence in such households (20–22) and gives rise to the co-occurrence of obesity and malnutrition (23,24). The historic aim of food aid interventions of any type is to provide food first, with the nutritional quality of the food a secondary consideration that may not support the specific needs of recipients in terms of diet and health (16,25,26). Given that much of the food is donated by the public, obtained through surplus food redistribution, or purchased by the providers, there is no guarantee that food from these sources will support a ‘healthy’ diet such as that recommended by the UK government. A recent systematic review demonstrates that the nutritional quality of food bank parcels is relatively poor, failing to meet nutritional requirements or cultural and health preferences (26).

Mental health can be negatively impacted by food insecurity and seeking food aid. Systematic reviews demonstrate that depression is strongly associated with food insecurity for parents (27), and for all adults food insecurity is a risk factor for stress or depression (18). Food insecurity is also shown to be associated with anxiety and depression during the Covid-19 pandemic (28). Qualitative studies suggest that worrying about food quality or having enough to eat as well as feeding children are some reasons this relationship between food insecurity and mental health is observed (29,30).

This paper reports the results from a mixed methods study in Wessex, South of England, which aimed to explore food insecurity, diet quality and mental wellbeing in participants using higher-agency food clubs. Pre and post intervention surveys are used to collect data along with semi-structured interviews with clients during 2022, the year in which the UK’s cost of living crisis was at its peak.

## Methods

### Food club settings

Two membership food clubs in Wessex were involved in this project. Both membership clubs offered a range of products including fresh fruit and vegetables; snacks (cakes, chocolate, biscuits); canned food, food cupboard staples (cereals, pasta, bread, rice); and household and personal hygiene items. The items were categorised into four or five groups and members selected a pre-specified number of items from each group. The groups were set by the clubs so could be different across the clubs but both clubs had a fruits and vegetables group in common. All members get the same quantity of food regardless of household size. Members at both clubs could additionally help themselves to “free food” items when available, which are foods that did not count towards their item limit but tended to be items available in surplus that could not be stored by the clubs, such as bread. Club A was associated with a food bank and had refrigeration facilities so offered meat, dairy products and frozen items including fruit and vegetables.

Club A was restricted to members from a specific geographic area and operated once a week for three hours at one of three locations. Club B allowed members to access any location multiple times a week, operating two different sites for two hours a week at each site. The opportunity to access at different locations meant that it was more accommodating to user’s schedules who could choose the alternate location or day if that suited them better. Both clubs offered hot drinks, social networking opportunities, and signposting to additional support/wraparound services. Club A also provided pastries from local outlets, invited representatives from various support organizations and at some sites, a “pay what you can” hot food option. During the study, two of Club A’s sites opened, while Club B experienced site changes (one closure), typical in food aid services.

In both clubs, members paid a fee to select items of food which were sourced mainly from FareShare (national network of surplus food redistribution to charities in the UK for use to support people in their communities; please see https://fareshare.org.uk/), other surplus donations or purchased from local supermarkets. At the start of the sample period most food originated from FareShare and surplus donations from local supermarkets, but as the cost-of-living crisis developed the volume of food purchased by the food clubs themselves increased and became the most common source of supply.

### Data collection

Participants aged 18 and over were recruited from the food clubs between March 31 and November 3, 2022. Members of the research team visited the sites, asked users to complete a brief survey, and invited them for an interview. Observations at the food clubs were recorded by the research team, including interactions between volunteers and service users and the way spaces were used in the clubs. Written informed consent was obtained.

*Surveys:* Participants completed a baseline survey at recruitment that collected data on diet, food practices, demographics, and health. Baseline data were collected retrospectively for participants at intervention sites that were already in place when the study commenced (one site each for club A and B) if participants had been service users for at least three months. These existing service users had used the interventions for less than 12 months at time of recruitment. New site users for the clubs which opened during the time of our study needed time to settle in prior to engaging in the study and were usually recruited at the second or third visit, two or three weeks after their initial visit to complete a baseline survey and then we followed up after three months from the date of their first survey.

Survey respondents who were already service users for at least three months completed the baseline and follow up surveys concurrently. We asked them to reflect on the time before they accessed the service and to answer the questions for this time period prior to using the food club for the baseline survey. Follow-up surveys included the same questions as the baseline survey (without demographics) and were completed after attending the clubs for at least three months. We discussed with our steering group and public representatives the implications of asking service users to complete a baseline survey retrospectively, where they had already been using the food club. There was agreement that for the questions asked in the survey, service users would have strong memories of their diet practices prior to using the service. Previous research has demonstrated that diet recall was reliable in retrospective surveys (31–34). However, we will focus on descriptive analysis.

We used the modified six-item USDA food security survey module (4) to ask about food security in the last 30 days and the short form Warwick-Edinburgh Mental Wellbeing Scale (WEMWBS) (35) to capture a measure of mental wellbeing. Food insecurity was calculated both using the USDA guidance and following the approach used by the Food Foundation in the UK (using responses to three of the six USDA module questions to capture moderate and severe experiences of food insecurity (36)). Diet quality was assessed using a food frequency questionnaire (FFQ) and the associated diet quality score, composed of fruit, vegetable, oily fish, fat and non-milk extrinsic sugar intakes, was calculated (37). The diet quality score ranged from 5 to 15 with higher scores indicating better diet quality. A question on the number of portions of fruit and vegetables consumed in a day was included, as well as the main cooking methods and barriers to a healthy diet. Participants received a £5 supermarket voucher as a thank you gesture for every survey completed.

Baseline and follow-up surveys were either completed on paper and subsequently entered into Qualtrics software by the research team or were completed by participants directly online using Qualtrics on personal devices (a QR code on advertising posters at club sites for baseline and email link for follow-up were provided).

*Interviews:* Participants who completed a baseline survey were invited to participate in a semi-structured interview on Microsoft Teams. Interviews explored pandemic experiences, their views on the food membership clubs and household eating habits. An interview guide was used for consistency and all interviews were conducted by NZ. Interviews lasted between 30 to 60 minutes and were recorded and transcribed using MS Teams transcription software. Transcripts were checked for accuracy against the recording and edited as appropriate to correct inaccuracies by NZ. Participants received a £20 supermarket voucher as a thank you gesture for their time.

### Data analysis

Statistical analysis was undertaken using Stata 17 (38). Descriptive percentages and summary statistics were generated for the full sample and for follow-up data.

Qualitative data was analysed using thematic analysis (39,40). Data analysis followed Braun and Clarke’s six-phase approach starting with familiarisation (reading and re-reading transcripts to note items of interest). Initial codes were generated by NZ with second coding of a sample of two interviews by DS using the codebook developed. No additional codes were identified on second coding. Based on the codes, themes were generated by reviewing the coded data to identify areas of similarity. Potential themes were reviewed in relation to the coded data and the entire dataset to ensure themes link to the coded data set. Themes were then defined and named; and finally the narrative of the data based on the analysis was constructed to produce the results summary.

### Stakeholder and Patient and Public Involvement (PPI)

A PPI contributor (FR) was involved in the design of the study, included as a co-applicant on the research funding proposal and consulted throughout the course of this study. The PPI contributor sat on the project steering group and contributed to the direction of the overall project, including development of the survey and interview topic guide, and data collection practices.

The steering group included stakeholders from local authorities and the food aid organisations. The survey was co-developed with steering group members to ensure data collected would be useful to inform decisions about the delivery of higher-agency food aid interventions. This included understanding barriers to household food security in this time- and resource-poor population, and contributed to questions in the interview guide. Surveys and interview guides used in data collection will be available as a toolkit for local authorities and food aid providers to use in their own evaluations. In April 2024, we returned to the food clubs to share the outcomes of this research with participants of the study and other food club members.

## Results

A total of 90 participants were recruited to the study; 56 from Club A and 34 from Club B. 52 participants completed the baseline and follow-up surveys. 38 out of the 52 participants with follow-up completed baseline and follow-up surveys at the same time as they had been attending the food club for at least three months at the time of recruitment. We will refer to these participants as the concurrent follow-up group hereafter.

75 participants were invited to interview as they included their contact information on the initial survey. Invitations were prioritised to reflect a distribution across the interventions and for households with children. In total 11 participants (21% of sample with follow-up) were interviewed. One interviewee attended both food clubs.

### Survey results

Characteristics of participants are shown in Table 1. 52% of participants at baseline were aged 35–54 years (n=43), 74% were female (n=63), 81% were of White ethnicity (n=73) and 71% reported having one or more dependent children (n=59).

**Table 1:** Characteristics of participants recruited from the food membership clubs.

|  | Baseline |  | Follow-up |  | Non-concurrent follow-up (at 3 months) |  | Concurrent follow-up |  |
| --- | --- | --- | --- | --- | --- | --- | --- | --- |
|  | n | % | n | % | n | % | n | % |
| Total n | 90 |  | 52 |  | 14 |  | 38 |  |
| Age, categorised |  |  |  |  |  |  |  |  |
| 18-24 | 4 | 4.8 | 2 | 4.4 | 0 | 0.0 | 2 | 6.3 |
| 25-34 | 14 | 16.9 | 6 | 13.3 | 3 | 23.1 | 3 | 9.4 |
| 35-44 | 26 | 31.3 | 16 | 35.6 | 5 | 38.5 | 11 | 34.4 |
| 45-54 | 17 | 20.5 | 10 | 22.2 | 4 | 30.8 | 6 | 18.8 |
| 55-64 | 13 | 15.7 | 6 | 13.3 | 0 | 0.0 | 6 | 18.8 |
| 65+ | 9 | 10.8 | 5 | 11.1 | 1 | 7.7 | 4 | 12.5 |
| Prefer not to say/no response | 7 | - | 7 | - | 1 | - | 6 | - |
| Gender |  |  |  |  |  |  |  |  |
| Female | 63 | 74.1 | 31 | 66.0 | 12 | 85.7 | 19 | 57.6 |
| Male | 22 | 25.9 | 16 | 34.0 | 2 | 14.3 | 14 | 42.4 |
| Prefer not to say/no response | 5 | - | 5 | - | - | - | 5 | - |
| Ethnicity |  |  |  |  |  |  |  |  |
| White | 73 | 81.1 | 39 | 75.0 | 14 | 100.0 | 25 | 65.8 |
| Asian | 10 | 11.1 | 8 | 15.4 | 0 | 0.0 | 8 | 21.1 |
| Mixed | 3 | 3.3 | 2 | 3.8 | 0 | 0.0 | 2 | 5.3 |
| Black | 3 | 3.3 | 3 | 5.8 | 0 | 0.0 | 3 | 7.9 |
| Other | 1 | 1.1 | 0 | 0.0 | 0 | 0.0 | 0 | 0.0 |
| Employment status |  |  |  |  |  |  |  |  |
| Employed full-time | 10 | 11.8 | 5 | 10.2 | 3 | 21.4 | 5 | 14.3 |
| Employed part-time | 17 | 20.0 | 9 | 18.4 | 0 | 0.0 | 6 | 17.1 |
| Unemployed | 45 | 52.9 | 27 | 55.1 | 6 | 42.9 | 21 | 60.0 |
| Self-employed | 4 | 4.7 | 4 | 8.2 | 3 | 21.4 | 1 | 2.9 |
| Retired | 4 | 4.7 | 2 | 4.1 | 1 | 7.1 | 1 | 2.9 |
| Student/trainee | 2 | 2.4 | 2 | 4.1 | 1 | 7.1 | 1 | 2.9 |
| Carer | 3 | 3.5 | 0 | 0.0 | 0 | 0.0 | 0 | 0.0 |
| Prefer not to say/no response | 5 | - | 3 | - | - | - | 3 | - |
| Household size |  |  |  |  |  |  |  |  |
| 1 | 19 | 21.1 | 11 | 21.2 | 0 | 0.0 | 11 | 28.9 |
| 2 | 16 | 17.8 | 13 | 25.0 | 2 | 14.3 | 11 | 28.9 |
| 3 | 16 | 17.8 | 11 | 21.2 | 4 | 28.6 | 7 | 18.4 |
| 4 | 17 | 18.9 | 8 | 15.4 | 4 | 28.6 | 4 | 10.5 |
| 5 or more | 22 | 24.4 | 9 | 17.3 | 4 | 28.6 | 5 | 13.2 |
| Dependent children in household |  |  |  |  |  |  |  |  |
| 0 | 24 | 28.9 | 14 | 29.8 | 1 | 7.1 | 13 | 34.2 |
| 1 | 16 | 19.3 | 11 | 23.4 | 3 | 21.4 | 8 | 21.1 |
| 2 | 23 | 27.7 | 13 | 27.7 | 7 | 50.0 | 6 | 15.8 |
| 3 or more | 20 | 24.1 | 9 | 19.1 | 3 | 21.4 | 6 | 15.8 |
| Prefer not to say/no response | 7 | - | 5 | - | - | - | 5 | 13.2 |
| Change in household size (stepchildren or other relatives sometimes stay) |  |  |  |  |  |  |  |  |
| Yes | 23 | 27.1 | 10 | 20.0 | 4 | 28.6 | 6 | 16.7 |
| Financial burden from change | 16 | 18.8 | 7 | 14.0 | 4 | 28.6 | 3 | 8.3 |
| No financial burden from change | 4 | 4.7 | 3 | 6.0 | 0 | 0.0 | 3 | 8.3 |
| Don't know | 1 | 1.2 | 0 | 0.0 | 0 | 0.0 | - | 0.0 |
| Prefer not to say | 2 | 2.4 | - | - | 0 | 0.0 | - | 0.0 |
| No | 62 | 72.9 | 40 | 80.0 | 10 | 71.4 | 30 | 83.3 |
| Prefer not to say/no response | 5 | - | 2 | - | - | - | 2 | - |
| Housing type |  |  |  |  |  |  |  |  |
| Owned/mortgaged | 19 | 24.7 | 7 | 16.3 | 3 | 25.0 | 4 | 12.9 |
| Social rented | 36 | 46.8 | 20 | 46.5 | 7 | 58.3 | 13 | 41.9 |
| Private rented | 16 | 20.8 | 13 | 30.2 | 1 | 8.3 | 12 | 38.7 |
| Other | 6 | 7.8 | 3 | 7.0 | 1 | 8.3 | 2 | 6.5 |
| Prefer not to say/no response | 13 | - | 9 | - | 2 | - | 7 | - |
| Registered disability |  |  |  |  |  |  |  |  |
| Yes | 20 | 25.3 | 12 | 25.5 | 4 | 30.8 | 8 | 23.5 |
| No | 59 | 74.7 | 35 | 74.5 | 9 | 69.2 | 26 | 76.5 |
| Prefer not to say/no response | 11 | - | 5 | - | 1 | - | 4 | - |

**Table 2:** Food insecurity, diet quality, mental wellbeing and food practices in the study sample with follow-up data (n=52), overall and by concurrency of follow-up.

|  | Baseline |  | Follow-up |  | Non-concurrent follow-up at 3 months (n=14) |  |  |  | Concurrent follow-up (n=38) |  |  |  |
| --- | --- | --- | --- | --- | --- | --- | --- | --- | --- | --- | --- | --- |
|  | Baseline |  | Follow-up |  | Baseline |  | Follow-up |  | Baseline |  | Follow-up |  |
|  | n | % | n | % | n | % | n | % | n | % | n | % |
| **Food insecurity** |  |  |  |  |  |  |  |  |  |  |  |  |
| Food insecurity status |  |  |  |  |  |  |  |  |  |  |  |  |
| High or marginal (food secure) | 4 | 10.3 | 16 | 41.0 | 2 | 22.2 | 1 | 11.1 | 2 | 5.3 | 15 | 50.0 |
| Low (food insecure) | 22 | 56.4 | 16 | 41.0 | 5 | 55.6 | 5 | 55.6 | 17 | 44.7 | 11 | 36.7 |
| Very low (food insecure) | 13 | 33.3 | 7 | 17.9 | 2 | 22.2 | 3 | 33.3 | 11 | 28.9 | 4 | 13.3 |
| Unable to categorise | 13 | - | 13 | - | 5 | - | 5 | - | 8 | - | 8 | - |
| Food security status (condensed using Food Foundation method) |  |  |  |  |  |  |  |  |  |  |  |  |
| Food secure | 14 | 26.9 | 30 | 60.0 | 2 | 14.3 | 4 | 30.8 | 12 | 31.6 | 26 | 70.3 |
| Food insecure | 38 | 73.1 | 20 | 40.0 | 12 | 85.7 | 9 | 69.2 | 26 | 68.4 | 11 | 29.7 |
| Unable to categorise | - | - | 2 | - | - | - | 1 | - | - | - | 1 | - |
| **Diet quality** |  |  |  |  |  |  |  |  |  |  |  |  |
| Diet quality score |  |  |  |  |  |  |  |  |  |  |  |  |
| 5-10 | 35 | 67.3 | 27 | 51.9 | 10 | 71.4 | 10 | 71.4 | 25 | 65.8 | 17 | 44.7 |
| 11-15 | 17 | 32.7 | 25 | 48.1 | 4 | 28.6 | 4 | 28.6 | 13 | 34.2 | 21 | 55.3 |
| Change in diet quality score |  |  |  |  |  |  |  |  |  |  |  |  |
| Decreased | - | - | 16 | 30.8 | - | - | 4 | 28.6 | - | - | 12 | 31.6 |
| No change | - | - | 13 | 25.0 | - | - | 5 | 35.7 | - | - | 8 | 21.1 |
| Increased | - | - | 23 | 44.2 | - | - | 5 | 35.7 | - | - | 18 | 47.4 |
| **The Warwick-Edinburgh Mental Wellbeing Scale (WEMWBS)** |  |  |  |  |  |  |  |  |  |  |  |  |
| Mental wellbeing |  |  |  |  |  |  |  |  |  |  |  |  |
| High | 0 | 0.0 | 4 | 7.8 | 0 | 0.0 | 0 | 0.0 | 0 | 0.0 | 4 | 10.8 |
| Average | 23 | 45.1 | 28 | 54.9 | 6 | 42.9 | 6 | 42.9 | 17 | 45.9 | 22 | 59.5 |
| Low | 28 | 54.9 | 19 | 37.3 | 8 | 57.1 | 8 | 57.1 | 20 | 54.1 | 11 | 29.7 |
| Unable to categorise | 1 | - | 1 | - | - | - | - | - | - | - | 1 | - |
| Change in mental wellbeing score |  |  |  |  |  |  |  |  |  |  |  |  |
| Decreased | - | - | 11 | 21.2 | - | - | 3 | 21.4 | - | - | 8 | 21.1 |
| No change | - | - | 10 | 19.2 | - | - | 4 | 28.6 | - | - | 6 | 15.8 |
| Increased | - | - | 31 | 59.6 | - | - | 7 | 50.0 | - | - | 24 | 63.2 |
| **Food practices** |  |  |  |  |  |  |  |  |  |  |  |  |
| I feel confident cooking |  |  |  |  |  |  |  |  |  |  |  |  |
| Strongly agree | 17 | 32.7 | 21 | 42.0 | 4 | 28.6 | 4 | 28.6 | 13 | 34.2 | 17 | 47.2 |
| Agree | 19 | 36.5 | 22 | 44.0 | 7 | 50.0 | 8 | 57.1 | 12 | 31.6 | 14 | 38.9 |
| Neither agree nor disagree | 13 | 25.0 | 6 | 12.0 | 3 | 21.4 | 2 | 14.3 | 10 | 26.3 | 4 | 11.1 |
| Disagree | 3 | 5.8 | 1 | 2.0 | - | - | - | - | 3 | 7.9 | 1 | 2.8 |
| Strongly disagree | 0 | 0.0 | 0 | 0.0 | - | - | - | - | - | - | - | - |
| No response | - | - | 2 | - | - | - | - | - | - | - | 2 | - |
| I cook lunch or the evening meal from scratch |  |  |  |  |  |  |  |  |  |  |  |  |
| Daily | 13 | 25.5 | 18 | 36.0 | - | - | 1 | 7.1 | 13 | 35.1 | 17 | 47.2 |
| 2-3 times a week | 19 | 37.3 | 20 | 40.0 | 7 | 50.0 | 8 | 57.1 | 12 | 32.4 | 12 | 33.3 |
| Weekly | 12 | 23.5 | 7 | 14.0 | 3 | 21.4 | 3 | 21.4 | 9 | 24.3 | 4 | 11.1 |
| Monthly | 4 | 7.8 | 2 | 4.0 | 3 | 21.4 | 1 | 7.1 | 1 | 2.7 | 1 | 2.8 |
| Less than once a month | 3 | 5.9 | 3 | 6.0 | 1 | 7.1 | 1 | 7.1 | 2 | 5.4 | 2 | 5.6 |
| No response | 1 | - | 2 | - | - | - | - | - | 1 | - | 2 | - |
| I plan meals before shopping |  |  |  |  |  |  |  |  |  |  |  |  |
| Almost always | 17 | 32.7 | 17 | 34.7 | 4 | 28.6 | 3 | 21.4 | 13 | 34.2 | 14 | 40.0 |
| Often | 3 | 5.8 | 10 | 20.4 | 2 | 14.3 | 2 | 14.3 | 1 | 2.6 | 8 | 22.9 |
| Sometimes | 19 | 36.5 | 10 | 20.4 | 4 | 28.6 | 4 | 28.6 | 15 | 39.5 | 6 | 17.1 |
| Rarely | 8 | 15.4 | 9 | 18.4 | 3 | 21.4 | 3 | 21.4 | 5 | 13.2 | 6 | 17.1 |
| Never | 5 | 9.6 | 3 | 6.1 | 1 | 7.1 | 2 | 14.3 | 4 | 10.5 | 1 | 2.9 |
| No response | - | - | 3 | - | - | - | - | - | - | - | 3 | - |
| I/we try new foods or recipes |  |  |  |  |  |  |  |  |  |  |  |  |
| Daily | 2 | 4.0 | 4 | 8.2 | - | - | - | - | 2 | 5.6 | 4 | 11.1 |
| 2-3 times a week | 3 | 6.0 | 8 | 16.3 | 2 | 14.3 | 1 | 7.7 | 1 | 2.8 | 7 | 19.4 |
| Weekly | 15 | 30.0 | 11 | 22.4 | 4 | 28.6 | 2 | 15.4 | 11 | 30.6 | 9 | 25.0 |
| Monthly | 10 | 20.0 | 14 | 28.6 | 2 | 14.3 | 5 | 38.5 | 8 | 22.2 | 9 | 25.0 |
| Less than once a month | 20 | 40.0 | 12 | 24.5 | 6 | 42.9 | 5 | 38.5 | 14 | 38.9 | 7 | 19.4 |
| No response | 2 | - | 3 | - | - | - | 1 | - | 2 | - | 2 | - |

### Food security

At baseline (recruitment), 58.5% of participants (n=51) reported skipping or cutting size of meals because there was not enough money for food (Supplementary Table 1). Among participants with follow-up data, 58% (n= 29) reported skipping or cutting size of meals at baseline (Figure 1). At follow-up, 69% (n=9) without concurrent follow-up and 15% (n=5) with concurrent follow-up reported skipping or cutting size of meals.

**Figure 1:**
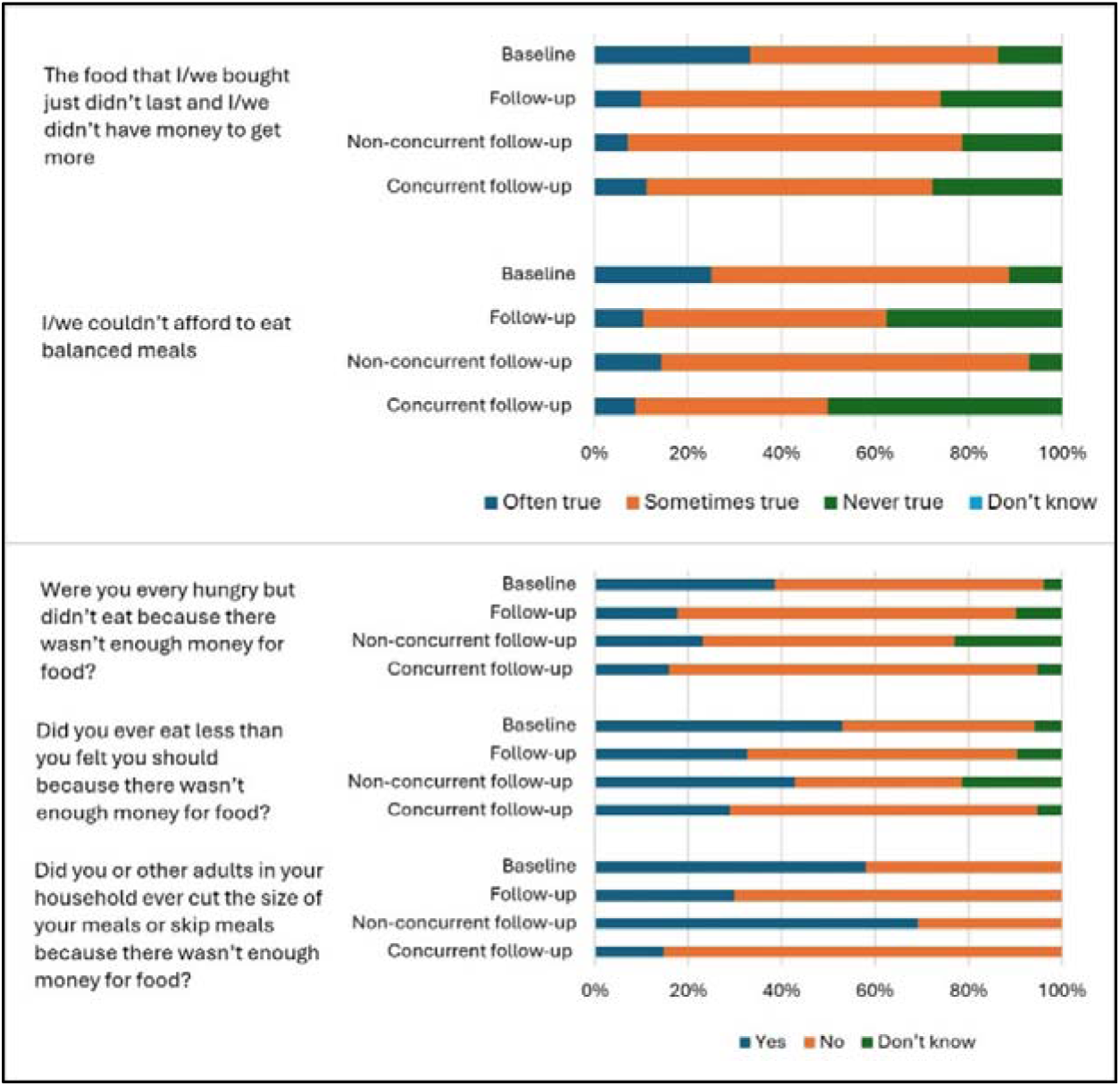
Responses to the food security module questions in the sample with follow-up (n=52) at baseline and follow-up.

**Figure 2:**
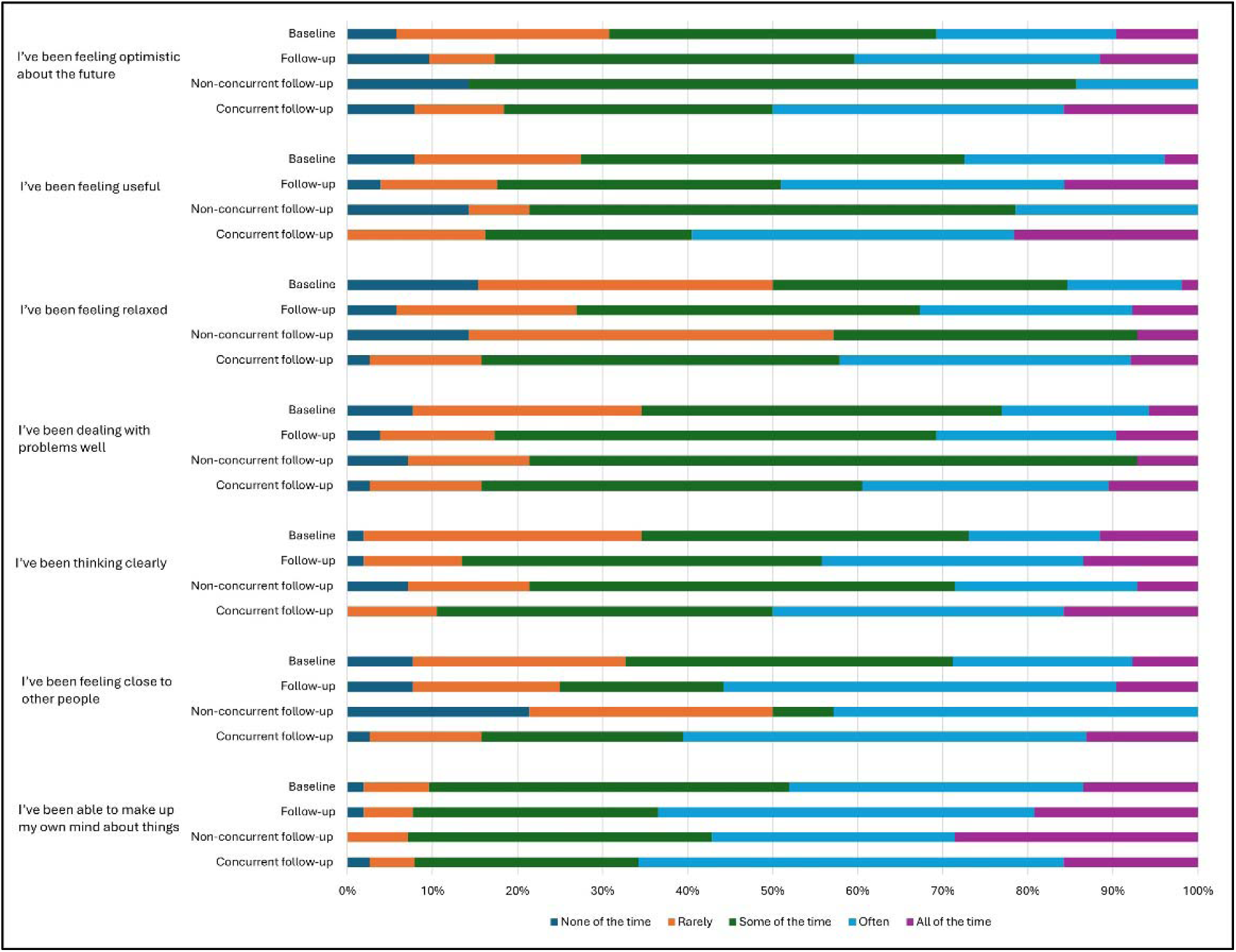
Responses to the short form Warwick-Edinburgh Mental Wellbeing Scale (WEMWBS) questions in the sample with follow-up (n=52) at baseline and follow-up.

Food security status at baseline was calculated for 69 participants who answered all questions of the USDA module, with 42% (n=29) reporting low food security and 43% (n=30) reporting very low food security.

Among participants with follow-up data, very low food security was 33% (n=13) at baseline, 33% (n=3) with non-concurrent follow-up and 13% (n=3) with concurrent follow-up. Low food security was 56% (n=22) at baseline, 55% (n=5) with non-concurrent follow-up and 37% (n=11) with concurrent follow-up. High or marginal food security was 10% (n=4) at baseline, 11% (n =1) with non-concurrent follow-up and 50% (n=15) with concurrent follow-up. Thirteen participants with follow-up data did not answer all six questions of the USDA food insecurity measure and thus we were unable to categorise their food insecurity status.

Using the Food Foundation approach, food insecurity was 68.4% (n=26) at baseline and 29.7% (n=11) at follow-up in participants with concurrent follow-up, and 85.7% (n=12) at baseline and 69.2% (n=9) at follow-up in participants who completed the surveys three months apart (non-concurrent follow-up).

### Diet quality

A third (32%, n=29) of 90 participants at baseline reported rarely or never eating fruit, with 23% (n=21) eating fruit at least once a day (Supplementary Table 2). For the sample with follow-up, 29% (n=15) at baseline, 36% (n=5) with non-concurrent follow-up and 5% (n=2) with concurrent follow-up reported rarely/never eating fruit at follow-up (Supplementary Tables 3 and 4). Similarly, 12% (n=6) at baseline, none with non-concurrent follow-up and 3% (n=1) with concurrent follow-up reported never eating vegetables. 23% (n=12) at baseline, 62% (n=8) with non-concurrent follow-up and 29% (n=11) had vegetables 2-3 times/week.

The proportion of participants with diet quality scores of 11 or more (better quality diet) was 32.7% (n=17) at baseline and 48.1% (n=6) at follow-up. In participants without concurrent follow-up, diet quality increased in 35.7% (median change 2, IQR 1 to 2), decreased in 28.6% (median change -1, IQR -1.5 to -1) and no change in 35.7% of participants. In participants with concurrent follow-up, diet quality increased in 47.4% (median change 1, IQR 1 to 2), decreased in 31.6% (median change -1, IQR -2 to -1) and no change in 21.1% of participants.

### Mental health

Based on scoring of responses to the WEMWBS questions, low wellbeing was 54.1% (n=20) at baseline and 29.7% (n=11) at follow-up in those with concurrent follow-up. Mental wellbeing increased in 50% of the sample (mean change 2.1 points, SD 1.2), decreased in 21.4% (mean change -6.4 points, SD 1.1) and no change in 28.6% of the sample without concurrent follow-up.

### Food practices

69.2% (n=36) participants reported feeling confident cooking at baseline and 86% (n=43) at follow-up). 10.1% (n=5) reported trying new foods or recipes more frequently at baseline, and 24.5% (n=12) at follow-up.

### Qualitative data: summary of main themes

Four of the interview participants completed the baseline and follow-up surveys concurrently whereas the remaining seven were users of newer sites and completed the surveys at baseline and after three months of using the interventions.

In this paper we focus on the most prominent themes from the data about impacts and experiences of food club. Additional themes included the impact of Covid, strategies for food access, trying new foods, accessibility and cost and adaptation due to cost of living which are less relevant for the aim of this paper.

## Impact of food club

Participants reported various positive impacts of food club on their diet, finances, health and social life.

### More varied diet

Fruit and vegetables are available at both food clubs and participants described eating more fruit and vegetables, some due to having access to these foods and some to partly avoid food waste. Participants described not wanting to waste food and thus making sure that they prioritised eating food that they thought was most likely to go off. Participants enjoyed the variety of fruit and vegetables available at the food club and reported trying to choose healthy food - *“because you have to get five options of fruit or veg every time, so it once it comes in the house. I don’t like wasting stuff now. So then I feel like I have to eat it so. Yes. So now I definitely, you know, make a real effort to eat the fruit or veg that comes. So I would say I have eat, I’m eating more of it now.” – P5”*

One participant described her frustration at previously having to buy biscuits as a snack for her children as it was the cheapest option available in the supermarket that would last a few days. She also described trying to ensure that her children have access to fruit (usually bananas and apples) and vegetables by shopping in budget supermarkets or buying reduced items but not being able to provide more variety of fruit. She went on to describe her joy at being able to get fruit from food club that they usually would not be able to buy from supermarkets more frequently due to the cost. This is in agreement with several participants who commented on diet improvement after using the food clubs - *“This has been a godsend [food club] cause it keeps us going, if that makes sense. We don’t run out now or cause I or I believe the children should have. Like I don’t have crisps but they should have as much unlimited fruit snacks as they want ….There’s certain foods that we wouldn’t normally have access…” – P8*

Some participants also described sometimes making different choices at food club compared to a supermarket. As food club is structured such that members can select a certain number of items from item groups, participants described selecting foods that they would not be willing to spend their money on in a supermarket. These were usually convenience or snack food such as crisps or sausage rolls which would normally not be part of their diet. Participants felt it was a balance of getting cheaper healthy food but also unhealthy food that may not necessarily form part of their diet if they had to pay for it in a supermarket - *“But they also had a lot of convenient food, like snacks. Hmm. Sausage rolls or crisps and all that. So some days you have to choose it because it’s on the options. So we also added lots of that type of snacks, which are not healthy. And I will probably not buy if I was paying with my money in the supermarket, but they were there and they were the option. So I chose them.” – P11*

### Less financial pressure

Food club took the financial strain off buying food to feed the family. Participants with bigger families reported having to do a shop in a supermarket, mostly in a budget supermarket or using points and vouchers from loyalty card schemes, partway through the week between club shops. Participants attending Club B also reported going to a supermarket for essentials not provided through the club, such as milk and other dairy products, due to the lack of refrigeration facilities in Club B.

Some participants reported being able to buy foods, commonly fresh fruit, in the supermarket that were generally deemed more expensive. Participants described the balance of getting food that was acceptable for members of the family at the food club (preferred fruit or at preferred ripeness/texture) meaning that they could then get the food which others preferred in the supermarket and feel less guilty or worry less about the cost. Some participants talked about wanting to eat healthier and lose weight, but the price attached to these choices was a barrier.

Participants also reported not buying certain foods, commonly fresh fruit or meat, as it was too expensive particularly with the current rates of inflation. They reported going without prior to going to food club or in between weekly food club visits once they had run out- *“obviously plenty of veg.. fruit and veg.. which I’ll get for my girls because they love their fruit and any other time it’ll probably won’t go and buy it in the shops of how much it is these days. It’s so expensive” – P7*

### Improved health, wellbeing and social interaction

Participants at Club A, which is restricted to members resident within a specific geographic area, reported walking to the food club quite frequently as the eligibility restriction meant that participants are more likely to live closer to the venue. Local participants at Club B also reported walking but others who lived further away had to travel by car or public transport to get to the club. Participants who walked there described it being nice to get out and get some fresh air.

Participants described a sense of relief after going to the food club as it gave them the opportunity to chat with other members and volunteers in addition to the food access which alleviated worries around feeding their family - *“I always feel happier after I’ve been there and I’ve had a chat and I’ve, I’ve sorted out my meals for the week and. Just a bit of relief.” – P6.* Some participants also felt that the interaction with members and volunteers helped their mental health, and they looked forward to going to the club and having the time to talk to others.

Participants described a warm welcoming atmosphere at the clubs. They found it nice to see the same people every week which led to new friendships and made them feel more integrated into the local community. Others also reported that the interactions at the club may be the only social interaction for them outside of their family that day making it something they looked forward to. The additional support services available at the club such as Citizens Advice or housing advice was welcomed as participants also felt they could easily get support with concerns or issues they were facing – “*There’s always something going on that’s the extra….So there’s kind of like a gateway. So though it’s just like a hub, isn’t it? You know there are and it’s not it’s… if you want it if that makes sense it’s there for you” – P8.* Members also shared their experiences in managing issues (such as around housing or bills) with each other and any tips they had learnt as part of their experience.

## Experiences of food club

### Recognising time and food range limitations

Although the experience of food club varied among participants, very few limitations were reported. The most common limitation was the need to get there early to ensure the trip was worthwhile so they had plenty of choices available but added to the time it took to get the shopping done. Some participants talked about the need to get there around half an hour to an hour before the food club opened to ensure they were early in the queue. Queuing outside before food club opened could be a pleasant experience in good weather but less so in wet and/or cold weather. This was sometimes a limiting factor for those with other commitments including education or part-time work and for those with health problems who could not stand in a queue outside for the necessary time - *“The only sort of downfall is if you don’t get there early, all the good stuff is gone. And I find now I’m going back into work. I just don’t have the time to spend…So yeah, yeah, it’s a shame if you’re if you’ve got no other commitments, then and you can spare two hours, then it’s fine.” – P1*.

Participants then had to queue inside when food club opened but this was generally described as a positive experience as participants had the opportunity to get a hot drink (in some clubs free pastries/biscuits) and socialise with the other members and volunteers. Participants who could not attend as frequently or who had not socialised as much at the food club said that it could feel a bit lonely, though generally everyone was friendly.

The other limitation that participants reported was around the range and quality of food. Only Club A provides refrigerated and frozen food so participants attending Club B had to source these products elsewhere. Most participants reported getting these items from a supermarket and that they did not mind doing so but others have chosen to visit other food clubs where these products are provided. As some of the food is sourced from surplus, participants mentioned having to check packaged fruit and vegetables as these could sometimes be a bit mouldy or bit wrinkly but was less easy for the staff at food club to identify due to being packaged.

### Sense of community

Participants described a sense of community and belonging from attending food club. The majority of participants lived locally to their club and felt they had made friends and connections locally that benefitted them in addition to food provision. Participants also described giving back to the food club through volunteering, donating spare produce that they had grown at home or in their allotment or through spreading the word about food club which lead to others donating their spare produce - *“But on the allotment, I do talk to a nice lady and she’s been donating some of her spare produce to the [food club]. So that’s been positive that I’ve been able to get a lot more people to give produce.” – P5*

### Overcoming stigma

Some participants reported being reluctant to go to food club or tell some people that they were going to food club and experiencing internalised stigma, which is when people accept negative associations to be true and applicable to themselves. The reluctance described was in communicating to people (other than close friends and family) that they were going to a food club - *“I think it’s not easy knowing you’re going to a food club. I usually tell people I’m going shopping. At the stigma attached? Maybe.” – P5*. Some participants described overcoming it after a while whereas others mentioned still working on overcoming it or choosing not to tell people. Some participants described being less worried about money or having food after attending food club for a while which helped overcome stigma – *“To start with, you know, I was a bit funny because of the stigma attached to it….But then after a while, I thought, you know what, I’m not really bothered… I found myself not having to (worry)… for any type of money to live or anything like that” – P3*.

However, participants were very clear that the staff and volunteers and the extended support community at food club were welcoming and did not make them feel judged or stigmatised.

## Discussion

There is a sense of a positive impact of food clubs on the food security, diet quality and mental wellbeing of participants when exploring both survey and interview data. The results are encouraging and reflect the range of experiences for clients of food clubs or similar aid schemes. A shift to higher consumption of fruit and vegetables after accessing the clubs was demonstrated strongly in the interviews. Participants spoke about the relief food aid offered in terms of cost of food and quality of diet, especially for their children. There was discussion as well of the stigma around accessing such support, but overall people who used these clubs spoke positively about the experience.

### Food security and diet quality in households

Among participants with follow-up, food security was 27% at baseline, 31% in those with non-concurrent follow-up and 70% in those with concurrent follow-up using the Food Foundation approach. Diet quality increased in 35.7% of participants without concurrent follow-up. This is consistent with a recent systematic review of 21 articles in high-income countries that noted improvements in diet quality in food bank users (likely due to their dominance as a form of food aid) (26). However, food parcels often failed to meet nutrient and individual requirements indicating that the quality of food parcels needed to improve, with greater provision of meat, fruit and vegetables alongside efforts to provide for cultural and health needs (26). Another systematic review of nine studies conducted in USA and Canada focussing on food aid use in households with children also found improvements in food security and diet quality, with models that provide choice and support services being most effective (41). Two studies in the USA that delivered a diabetes prevention intervention to food bank users at risk of diabetes highlighted improvements in food security and fruit and vegetable consumption (42,43). A longitudinal study in Canada contrasting food banks with a choice model (similar to the food clubs studied here) also demonstrated greater improvements to household food security in those using food banks with a choice model compared to those using food banks without a choice model (12). Our positive findings were reflected in interviews across the themes of diet variety and financial pressure, with participants also reporting higher fruit and vegetable consumption in the diet questions.

It must be noted that food insecurity is a challenging concept to capture in survey questions. We chose to ask participants to reflect on the previous 30 days when answering the questions about food insecurity. We note, however, that some people may experience food insecurity for brief periods of time due to a sudden change in circumstance (job loss, relationship breakdown or unexpected costs) or they may experience this over a longer period of time due to longer term ill health or needing to care for dependents, or an accumulation of challenging circumstances (25,44). The increased cost of living in the UK during the study period has meant that the intended aim of food aid to be a temporary source of support has become a more longer term feature of the food environment. During the period of data collection, food prices were increasing for many basic items, which will also impact on household food security as budgets were further stretched (5).

The food clubs we worked with in this study intended their use to be temporary, in the case of Club A they wanted members to move on to purchasing all of their food in retail outlets after a year. However, this was not feasible in practice. When we returned to share the results of the research with food club members, we recognised participants from more than a year prior. Discussion of the research outcomes with the research team highlighted that for those still using the clubs, they did not have enough money to purchase all of their food from standard retail options. Some of the differences in outcomes observed between those who concurrently completed the first two surveys and those who did not may be accounted for because those who had already been using the food club for longer periods of time were in a more entrenched time of food insecurity, and there would need to be a more substantial change to their circumstances to be reflected in food security questions in a survey.

Fruit and vegetable provision is a key area for both food clubs included in this study and all interview participants described household members consuming more fruit and vegetables. However, the extent of diet changes may not be well captured in the survey results as these reflect the diet quality of one household member participating in the study. In the interviews participants told us that they were eating and feeling better since going to food club. Interview participants with children told us that the fruit tended to be more frequently consumed by the children after using the food club, as well as vegetables such as carrots which are frequently consumed raw as a snack.

Sometimes there were less familiar vegetables available at food club. Details were given about how to cook them, or suggested recipes. Participants liked this learning aspect of food club which meant they tried new foods but could also have the option of providing a more familiar meal to members of the household who were hesitant to try new foods. Other participants described being able to buy preferred fruit for a household member when they went to a supermarket for a usual food shop. Without the food club, they could not afford these items, which would have meant that household member would not have any fruit in their diet. As some of the food available at food club have longer shelf lives (such as cereal, dried pasta, canned beans), participants described the sense of relief knowing that there was food in the cupboard and thus some meals for the future were in place.

Overall, in both the surveys and interviews we can see a promising trajectory for diet quality among those using the food clubs. In future surveys, researchers need to account for length of time a participant has been using the clubs and note that surveys are only capturing the experience of one person through the survey. We know that parents reduce their food intake to shield children from food insecurity (45), so any intervention may take longer to observe any positive change for parents. Further, there were a substantial number of cases where we could not categorise food security status using the standard USDA survey responses as people skipped the question about how many days they had skipped meals. When we used the same approach as the Food Foundation, we were able to classify almost all respondents in terms of food security status.

A relatively complex FFQ may not be most effective at identifying diet change as some practices may not change, such as consumption of fish, due to an overall low food budget and household diet preferences. Fruit and vegetable consumption is a useful measure, as this is something participants discussed unprompted, and it is a well-known healthy diet aim. Food costs remained high and increased significantly during the study period (5), so clubs may not have as much of a measurable impact as in more affluent times.

### Mental wellbeing and community

Wellbeing or mental health are other notable concerns in food-insecure households, and mental wellbeing increased in 50% of the sample with non-concurrent follow-up and in 63% with concurrent follow-up. Comments made during the interviews pointed to reduced worry over food and financial concerns, improved diet options and the engagement with other clients and volunteers as the reasons for enhanced mental wellbeing. The improvement observed in our work is in agreement with results from Canada after an 18 month follow-up (12). The Canadian study was larger and recruited participants accessing different types of food banks (standard food bank parcel, food bank with choices (similar to food clubs in this study) and food banks with additional onsite services) with follow-up at 6, 12 and 18 months and found improvement in mental health at the 18-month follow-up point. A recent study in the UK explored social impacts of a wide range of aid including food club/pantry, food banks and community kitchens. Researchers interviewed coordinators and other representatives of aid and demonstrated the positive impacts of food hubs (including food clubs) for individual wellbeing of clients, suggesting this was the greatest positive impact they observed (46).

Participants who agreed to the interview told us about looking forward to the day they would go to food club, not only because they could get food (relieving financial stress) but to be able to interact with other users, staff and volunteers, all of which improved their mental health. Participants who lived close to the clubs talked about walking to the club when the weather was good and how this helped their mental and physical health.

Participants in our study spoke about the negative aspect of stigma in accessing food aid, but noted that this also improved as time passed, potentially due to the welcoming environment at the clubs, interacting with other people in a similar situation and seeing the same people every week enabling a sense of community. Overall, there were many positive reflections on the ability to talk with others; building a community for wider social support were central, as observed in other studies of food clubs (47). This role, as a place to build community, is echoed in the report on affordable food clubs by Feeding Britain (48).

### Strengths and limitations

There are several strengths to this work. The survey tool and interview guide were developed with several local stakeholders and public input, to ensure the data collected will inform practice in the food aid settings. The combination of survey data and interviews with participants allowed us to explore the possible reasons behind patterns noted from the surveys. There is limited literature on the impacts of food aid in forms other than food banks, and here we present a mixed-methods study which allowed us to follow-up with participants as they settle into the intervention. Using validated measures for food security and wellbeing provide comparability for future studies into the impact of higher-agency food aid.

There are some limitations to this study. Some food clubs were established when we recruited participants, and most participants completed the baseline and follow up surveys together. Both the USDA food insecurity module and FFQs collect data over the previous 12 months so the time lag is established in the data capture instruments but the risk of measurement error remains. We have presented results for the overall follow-up as well as by participants with concurrent follow-up or not for transparency though the sample size by this categorisation is small. There also is a risk of social desirability bias for both survey and interview responses. Our sample was predominantly female and of White ethnicity and therefore the findings may not be generalisable to other groups.

### Recommendations for future research and policy

Although FFQs are validated tools, they can be complex to complete for participants. For example we found some confusion regarding the unit of measurement (which ranged from portions per day to portions per week). In addition, people may be less familiar with some food descriptions such as beans or pulses, or differentiating between fibre rich and non-fibre rich breakfast cereal. Simple data collection tools like describing a portion size of fruit and vegetables then asking about portion consumption, and the length of surveys should be considered when designing studies. Similarly, we were unable to categorise food insecurity for about a quarter of responses using the USDA measure. This was mainly because participants struggled with the question on how many days they skipped meals. Using the Food Foundation approach of using three questions out of the six-item USDA measure, we were able to categorise food insecurity for almost all participants at both time points (two responses could not be categorised). The Food Foundation approach also aims to capture more moderate and severe experiences of food insecurity (36).

Giving newer club users time to settle-in prior to recruitment to the surveys or interviews worked well in this study. We attended food clubs regularly during the recruitment period so users became familiar with us and were used to having us around. We interacted with new users when they were willing to and mentioned the study but waited until the second or third visit/week to recruit. This worked well for this study as we found that the new members’ first visit to food club was quite overwhelming for most people, and they needed some time to familiarise themselves. Follow-up can be difficult in this sample as most people did not respond to phone calls, text messages or emails (49). The stakeholders informed us that follow-up would be difficult due to difficulties experienced in previous attempts to contact users. We did not capture data on the food that participants purchased from food club or compare the differences between foods available at the two clubs and this could potentially be important work to inform future food provision.

In terms of policy, we agree that for optimal flexibility and dignity, cash-first response to food insecurity is best as noted by others (8). However, as one of our coordinators of food aid stated, “Food aid is where we are right now.” In this context we aim to improve the process of collecting insight about the outcomes for people accessing food aid, using simplified surveys and interviews. To further collect meaningful data, we will further engage with people who have lived experience of food insecurity to work in and with these communities.

Ideally there will be extended state intervention to support food insecure households as the welfare system has yet to fully meet this need through increasing income via benefit payments, Healthy Start vouchers or similar. We observed over the time of this study a decline in the amount of surplus food available to the food clubs, and more food had to be purchased to meet demand. The reduction in surplus food is positive in terms of less food potentially going to waste, however, this demonstrates the precarious balance of food aid. The main policy response to household food insecurity is food aid but the sector relies predominantly on donations or support from local government (5). There is an irony to see a sector which has developed to respond to households facing their own precarity in terms of food supply experiencing similar challenges of sufficient food and staffing, where often volunteers represent a substantial portion of their workforce (44). There are positive moves to improve the quality and variety of food available through food aid in any format (5,16). Access to aid can be improved through more non-referral options, and longer opening hours clubs including weekends or after usual working hours.

### Conclusions

This novel exploration of food aid highlights the high prevalence of food insecurity in those accessing food clubs in Wessex and demonstrates improvements in diet and mental wellbeing and decreases in food insecurity after accessing food clubs. Food aid is changing as there is recognition of the need to provide a more sustainable and supportive model to develop food security. The shift from food banks to higher-agency food aid, such as food clubs, provides an opportunity to refine the delivery of interventions to optimise outcomes for the populations they serve, in terms of products available, recipe cards, or wraparound services available for signposting. Collecting data from clients of these services which allows comparisons over time through standardised survey modules adds valuable data about the potential benefits of food clubs/pantries. Understanding more about the experiences of clients through interviews offers new insights regarding the perceived value and opportunities for improvement in supporting diet quality and wellbeing. Uniting these methods provides a process for ongoing data collection and reflection of the relative success of food clubs until we move to a point where greater equity is achieved, and such aid is no longer embedded in the food environment.

## List of abbreviations

USDA : United States Department of Agriculture

WEMWBS: Warwick-Edinburgh Mental Wellbeing Scale

## Declarations

## Ethics approval and consent to participate

Ethical approval was granted by the University of Southampton Faculty of Environmental and Life Sciences committee (ERGO 68876). Informed consent was obtained from all study participants.

## Consent for publication

Not applicable.

## Availability of data and materials

The anonymised datasets analysed during the current study may be available from the corresponding authors on reasonable request, pending approval from the relevant ethics committee and research governance structures.

## Competing interests

BM is a trustee of Southampton Social Aid Group. All other authors have no conflicts of interest to declare.

## Funding

This work was supported by the National Institute for Health Research (NIHR) Applied Research Collaboration (ARC) Wessex. The funder had no role in study design, data collection and analysis, decision to publish, or preparation of the manuscript. The views expressed in this paper are those of the authors and not necessarily those of the National Institute of Health and Care Research or the Department for Health and Social Care. For the purpose of open access, the author has applied a Creative Commons Attribution (CC BY 4.0) licence to any Author Accepted Manuscript version arising.

## Authors’ contributions

Authors contributions were as follows: study concept (NAA, DS), study methodology (all authors), data collection (NZ, ET, NAA, DS,), data analysis (NZ, ET), drafting of the manuscript (NZ, DS), and revising for content (all authors). All authors read and approved the final manuscript.

## Acknowledgements

We would like to thank the study participants, and our PPI representatives for their time and invaluable contribution to this work. We would like to thank all the staff and volunteers at the food membership clubs for facilitating study recruitment.

## Authors’ information

NZ and ET made equal contribution as first author.

## Footnotes

1 In receipt of Universal credit with household income less than £7400/year or other benefits.

## Notes

### Author Declarations

The Faculty of Environmental and Life Sciences committee (ERGO 68876) of University of Southampton gave ethical approval for this work.

### Summary of Updates

This version of the manuscript has been revised based on peer review comments.

## References

1. Loopstra R. Interventions to address household food insecurity in high-income countries. Proc Nutr Soc. 2018 Aug;77(3):270–81.

2. Dixon LB, Winkleby MA, Radimer KL. Dietary Intakes and Serum Nutrients Differ between Adults from Food-Insufficient and Food-Sufficient Families: Third National Health and Nutrition Examination Survey, 1988–1994. The Journal of Nutrition. 2001 Apr;131(4):1232–46.

3. Kirkpatrick SI, Tarasuk V. Food Insecurity Is Associated with Nutrient Inadequacies among Canadian Adults and Adolescents3. The Journal of Nutrition. 2008 Mar;138(3):604–12.

4. United States Department of Agriculture. U.S. Household Food Security Survey Module [Internet]. Available from: https://www.ers.usda.gov/topics/food-nutrition-assistance/food-security-in-the-u-s/survey-tools/#six

5. Department for Environment, Food & Rural Affairs. United Kingdom Food Security Report 2024: Theme 4: Food Security at Household Level [Internet]. 2024. Available from: https://www.gov.uk/government/statistics/united-kingdom-food-security-report-2024/united-kingdom-food-security-report-2024-theme-4-food-security-at-household-level#introduction

6. Taylor N, Boyland E, Hardman CA. Conceptualising food banking in the UK from drivers of use to impacts on health and wellbeing: A systematic review and directed content analysis. Appetite. 2024 Dec;203:107699.

7. Dowler E, Lambie-Mumford H. How Can Households Eat in austerity? Challenges for Social Policy in the UK. Social Policy & Society. 2015 Jul;14(3):417–28.

8. Milbourne P. Beyond ‘feeding the crisis’: Mobilising ‘more than food aid’ approaches to food poverty in the UK. Geoforum. 2024 Mar;150:103976.

9. Thompson C, Smith D, Cummins S. Understanding the health and wellbeing challenges of the food banking system: A qualitative study of food bank users, providers and referrers in London. Social Science & Medicine. 2018 Aug;211:95–101.

10. Douglas F, Sapko J, Kiezebrink K, Kyle J. Resourcefulness, Desperation, Shame, Gratitude and Powerlessness: Common Themes Emerging from A Study of Food Bank Use in Northeast Scotland. AIMS Public Health. 2015;2(3):297–317.

11. Garthwaite K. Stigma, shame and ‘people like us’: an ethnographic study of foodbank use in the UK. Journal of Poverty and Social Justice. 2016 Oct;24(3):277–89.

12. Rizvi A, Wasfi R, Enns A, Kristjansson E. The impact of novel and traditional food bank approaches on food insecurity: a longitudinal study in Ottawa, Canada. BMC Public Health. 2021 Dec;21(1):771.

13. Jones CL, Ksobiech K, Maclin K. “They Do a Wonderful Job of Surviving”: Supportive Communication Exchanges Between Volunteers and Users of a Choice Food Pantry. Journal of Hunger & Environmental Nutrition. 2019 Mar 4;14(1–2):204–24.

14. Martin KS, Wu R, Wolff M, Colantonio AG, Grady J. A Novel Food Pantry Program. American Journal of Preventive Medicine. 2013 Nov;45(5):569–75.

15. Mukoya MN, McKay FH, Dunn M. Can Giving Clients a Choice in Food Selection Help to Meet Their Nutritional Needs?: Investigating a Novel Food Bank Approach for Asylum Seekers. Int Migration & Integration. 2017 Nov;18(4):981–91.

16. Williams A, May J. A genealogy of the food bank: Historicising the rise of food charity in the UK. Trans Inst British Geog. 2022 Sep;47(3):618–34.

17. Thompson C, Smith D, Cummins S. Food banking and emergency food aid: expanding the definition of local food environments and systems. Int J Behav Nutr Phys Act. 2019 Jan 7;16(1):2.

18. Pourmotabbed A, Moradi S, Babaei A, Ghavami A, Mohammadi H, Jalili C, et al. Food insecurity and mental health: a systematic review and meta-analysis. Public Health Nutr. 2020 Jul;23(10):1778–90.

19. Monsivais P, Mclain J, Drewnowski A. The rising disparity in the price of healthful foods: 2004–2008. Food Policy. 2010 Dec;35(6):514–20.

20. Hanson KL, Connor LM. Food insecurity and dietary quality in US adults and children: a systematic review. The American Journal of Clinical Nutrition. 2014 Aug;100(2):684–92.

21. Shinwell J, Defeyter MA. Food Insecurity: A Constant Factor in the Lives of Low-Income Families in Scotland and England. Front Public Health. 2021 May 19;9:588254.

22. Armstrong B, Hepworth AD, Black MM. Hunger in the household: Food insecurity and associations with maternal eating and toddler feeding. Pediatric Obesity. 2020 Oct;15(10):e12637.

23. Crawford PB, Webb KL. Unraveling the Paradox of Concurrent Food Insecurity and Obesity. American Journal of Preventive Medicine. 2011 Feb;40(2):274–5.

24. Drewnowski A, Specter S. Poverty and obesity: the role of energy density and energy costs. The American Journal of Clinical Nutrition. 2004 Jan;79(1):6–16.

25. Garthwaite KA, Collins PJ, Bambra C. Food for thought: An ethnographic study of negotiating ill health and food insecurity in a UK foodbank. Social Science & Medicine. 2015 May;132:38–44.

26. Oldroyd L, Eskandari F, Pratt C, Lake AA. The nutritional quality of food parcels provided by food banks and the effectiveness of food banks at reducing food insecurity in developed countries: a mixed-method systematic review. J Human Nutrition Diet. 2022 Dec;35(6):1202–29.

27. Cain KS, Meyer SC, Cummer E, Patel KK, Casacchia NJ, Montez K, et al. Association of Food Insecurity with Mental Health Outcomes in Parents and Children. Academic Pediatrics. 2022 Sep;22(7):1105–14.

28. Cai J, Parker M, Tekwe C, Bidulescu A. Food insecurity and mental health among US adults during the COVID-19 pandemic: Results from National Health Interview Survey, 2020–2021. Journal of Affective Disorders. 2024 Jul;356:707–14.

29. Liebe RA, Porter KJ, Adams LM, Hedrick VE, Serrano EL, Cook N, et al. “I’m Doing the Best that I Can”: Mothers Lived Experience with Food Insecurity, Coping Strategies, and Mental Health Implications. Current Developments in Nutrition. 2024 Apr;8(4):102136.

30. Lindow P, Yen IH, Xiao M, Leung CW. ‘You run out of hope’: an exploration of low-income parents’ experiences with food insecurity using Photovoice. Public Health Nutr. 2022 Apr;25(4):987–93.

31. Egele VS, Klopp E, Stark R. Evaluating self-reported retrospective average daily fruit, vegetable, and egg intake: Trustworthy—Sometimes! Applied Psych Health & Well. 2023 Aug;15(3):1130–49.

32. Rohan TE, Potter JD. RETROSPECTIVE ASSESSMENT OF DIETARY INTAKE. American Journal of Epidemiology. 1984 Dec 1;120(6):876–87.

33. Schmidt RJ, Goodrich AJ, Granillo L, Huang Y, Krakowiak P, Widaman A, et al. Reliability of a short diet and vitamin supplement questionnaire for retrospective collection of maternal nutrient intake. Global Epidemiology. 2024 Dec;8:100150.

34. Raidl M, Johnson S, Gardiner K, Denham M, Spain K, Lanting R, et al. Use Retrospective Surveys to Obtain Complete Data Sets and Measure Impact in Extension Programs. Journal of Extension. 2004 Apr;42(2):2RIB2.

35. Tennant R, Hiller L, Fishwick R, Platt S, Joseph S, Weich S, et al. The Warwick-Edinburgh Mental Well-being Scale (WEMWBS): development and UK validation. Health Qual Life Outcomes. 2007 Dec;5(1):63.

36. The Food Foundation. Food insecurity tracking [Internet]. 2024 [cited 2024 Sep 14]. Available from: https://foodfoundation.org.uk/initiatives/food-insecurity-tracking

37. Cleghorn CL, Harrison RA, Ransley JK, Wilkinson S, Thomas J, Cade JE. Can a dietary quality score derived from a short-form FFQ assess dietary quality in UK adult population surveys? Public Health Nutr. 2016 Nov;19(16):2915–23.

38. Stata Statistical Software: Release 17. College Station, TX: StataCorp LLC; 2021.

39. Braun V, Clarke V. Using thematic analysis in psychology. Qualitative Research in Psychology. 2006 Jan;3(2):77–101.

40. Braun V, Clarke V. Thematic analysis. In: Cooper H, Camic PM, Long DL, Panter AT, Rindskopf D, Sher KJ, editors. APA handbook of research methods in psychology, Vol 2: Research designs: Quantitative, qualitative, neuropsychological, and biological [Internet]. Washington: American Psychological Association; 2012 [cited 2024 Nov 14]. p. 57–71. Available from: https://content.apa.org/books/13620-004

41. Stahacz C, Alwan NA, Taylor E, Smith D, Ziauddeen N. The impact of food aid interventions on food insecurity, diet quality and mental health in households with children in high-income countries: a systematic review. Public Health Nutr. 2024;27(1):e195.

42. Seligman HK, Smith M, Rosenmoss S, Marshall MB, Waxman E. Comprehensive Diabetes Self-Management Support From Food Banks: A Randomized Controlled Trial. Am J Public Health. 2018 Sep;108(9):1227–34.

43. Cheyne K, Smith M, Felter EM, Orozco M, Steiner EA, Park Y, et al. Food Bank-Based Diabetes Prevention Intervention to Address Food Security, Dietary Intake, and Physical Activity in a Food-Insecure Cohort at High Risk for Diabetes. Prev Chronic Dis. 2020 Jan 9;17:E04.

44. Smith D, Thompson C. Food Deserts and Food Insecurity in the UK: Exploring Social Inequality [Internet]. 1st ed. London: Routledge; 2022 [cited 2025 Feb 3]. Available from: https://www.taylorfrancis.com/books/9781003184560

45. Ovenell M, Azevedo Da Silva M, Elgar FJ. Shielding children from food insecurity and its association with mental health and well-being in Canadian households. Can J Public Health. 2022 Apr;113(2):250–9.

46. Papargyropoulou E, Bridge G, Woodcock S, Strachan E, Rowlands J, Boniface E. Impact of food hubs on food security and sustainability: Food hubs perspectives from Leeds, UK. Food Policy. 2024 Oct;128:102705.

47. Moraes C, McEachern MG, Gibbons A, Scullion L. Understanding Lived Experiences of Food Insecurity through a Paraliminality Lens. Sociology. 2021 Dec;55(6):1169–90.

48. Feeding Britain. Affordable Food Clubs Impact Report (April 2024) [Internet]. Available from: https://feedingbritain.org/affordable-food-clubs-impact-report-april-2024/

49. Bonevski B, Randell M, Paul C, Chapman K, Twyman L, Bryant J, et al. Reaching the hard-to-reach: a systematic review of strategies for improving health and medical research with socially disadvantaged groups. BMC Med Res Methodol. 2014 Dec;14(1):42.

