## Supplementary Table for "A mixed methods study exploring food insecurity and diet quality in households accessing food clubs in England"

Supplementary Table 1: Food insecurity, diet quality, mental wellbeing and food practices at baseline in the study sample

|  | n | % |
| --- | --- | --- |
| Total n | 90 |  |
| **Food insecurity questions** |  |  |
| The food that I/we bought just didn’t last and I/we didn’t have money to get more |  |  |
| Often true | 26 | 29.9 |
| Sometimes true | 47 | 54.0 |
| Never true | 14 | 16.1 |
| Don’t know | 0 | 0.0 |
| No response | 3 | - |
| I/we couldn’t afford to eat balanced meals |  |  |
| Often true | 21 | 26.6 |
| Sometimes true | 44 | 55.7 |
| Never true | 14 | 17.7 |
| Don’t know | 0 | 0.0 |
| No response | 11 | - |
| Did you or other adults in your household ever cut the size of your meals or skip meals because there wasn’t enough money for food? |  |  |
| Yes | 51 | 58.0 |
| No | 37 | 42.0 |
| Don’t know | 0 | 0.0 |
| No response | 2 | - |
| Did you ever eat less than you felt you should because there wasn’t enough money for food? |  |  |
| Yes | 50 | 56.2 |
| No | 34 | 38.2 |
| Don’t know | 5 | 5.6 |
| No response | 1 | - |
| Were you every hungry but didn’t eat because there wasn’t enough money for food? |  |  |
| Yes | 39 | 43.8 |
| No | 47 | 52.8 |
| Don’t know | 3 | 3.4 |
| No response | 1 | - |
| Food security status |  |  |
| High or marginal (food secure) | 10 | 14.5 |
| Low (food insecure) | 29 | 42.0 |
| Very low (food insecure) | 30 | 43.5 |
| Unable to categorise | 21 | - |
| Food security status (condensed using Food Foundation method) |  |  |
| Food secure | 25 | 27.8 |
| Food insecure | 65 | 72.2 |
| **Diet quality** |  |  |
| Diet quality score |  |  |
| 5-10 | 63 | 70.0 |
| 11-15 | 27 | 30.0 |
| [**The Warwick-Edinburgh Mental Wellbeing Scale (WEMWBS)**](https://warwick.ac.uk/fac/sci/med/research/platform/wemwbs/) |  |  |
| I’ve been feeling optimistic about the future |  |  |
| None of the time | 6 | 6.7 |
| Rarely | 22 | 24.7 |
| Some of the time | 30 | 33.7 |
| Often | 20 | 22.5 |
| All of the time | 11 | 12.4 |
| No response | 1 | - |
| I’ve been feeling useful |  |  |
| None of the time | 7 | 8.0 |
| Rarely | 23 | 26.4 |
| Some of the time | 34 | 39.1 |
| Often | 19 | 21.8 |
| All of the time | 4 | 4.6 |
| No response | 3 | - |
| I’ve been feeling relaxed |  |  |
| None of the time | 18 | 20.5 |
| Rarely | 26 | 29.5 |
| Some of the time | 27 | 30.7 |
| Often | 14 | 15.9 |
| All of the time | 3 | 3.4 |
| No response | 2 | - |
| I’ve been dealing with problems well |  |  |
| None of the time | 8 | 9.2 |
| Rarely | 20 | 23.0 |
| Some of the time | 37 | 42.5 |
| Often | 17 | 19.5 |
| All of the time | 5 | 5.7 |
| No response | 3 | - |
| I’ve been thinking clearly |  |  |
| None of the time | 5 | 5.8 |
| Rarely | 24 | 27.9 |
| Some of the time | 36 | 41.9 |
| Often | 14 | 16.3 |
| All of the time | 7 | 8.1 |
| No response | 4 | - |
| I’ve been feeling close to other people |  |  |
| None of the time | 6 | 6.8 |
| Rarely | 22 | 25.0 |
| Some of the time | 32 | 36.4 |
| Often | 20 | 22.7 |
| All of the time | 8 | 9.1 |
| No response | 2 | - |
| I’ve been able to make up my own mind about things |  |  |
| None of the time | 3 | 3.4 |
| Rarely | 10 | 11.4 |
| Some of the time | 32 | 36.4 |
| Often | 30 | 34.1 |
| All of the time | 13 | 14.8 |
| No response | 2 | - |
| Mental wellbeing |  |  |
| High | 1 | 1.2 |
| Average | 40 | 47.1 |
| Low | 44 | 51.8 |
| Unable to categorise | 5 | - |
| **Food practices** |  |  |
| I feel confident cooking |  |  |
| Strongly agree | 34 | 38.6 |
| Agree | 31 | 35.2 |
| Neither agree nor disagree | 19 | 21.6 |
| Disagree | 3 | 3.4 |
| Strongly disagree | 1 | 1.1 |
| No response | 2 | - |
| I cook lunch or the evening meal from scratch |  |  |
| Daily | 23 | 26.1 |
| 2-3 times a week | 39 | 44.3 |
| Weekly | 15 | 17.0 |
| Monthly | 5 | 5.7 |
| Less than once a month | 6 | 6.8 |
| No response | 2 | - |
| I plan meals before shopping |  |  |
| Almost always | 27 | 30.3 |
| Often | 10 | 11.2 |
| Sometimes | 30 | 33.7 |
| Rarely | 16 | 18.0 |
| Never | 6 | 6.7 |
| No response | 1 | - |
| I/we try new foods or recipes |  |  |
| Daily | 3 | 3.4 |
| 2-3 times a week | 10 | 11.5 |
| Weekly | 26 | 29.9 |
| Monthly | 18 | 20.7 |
| Less than once a month | 30 | 34.5 |
| No response | 3 | - |

Supplementary Table 2: Responses to food frequency questionnaire items (in percentages) at baseline in the study sample (n=90)

|  | Rarely/Never | < once a week | Once a week | 2-3 times a week | 4-6 times a week | 1-2 times a day | 3-4 times a day | 5+ times a day | No response |
| --- | --- | --- | --- | --- | --- | --- | --- | --- | --- |
| Fruit (tinned/fresh) | 32 | 10 | 8 | 20 | 7 | 13 | 6 | 4 | 0 |
| Fruit juice (not cordial or squash) | 47 | 20 | 16 | 7 | 4 | 6 | 0 | 0 | 1 |
| Salad (not garnish added to sandwiches) | 24 | 18 | 19 | 22 | 7 | 9 | 0 | 0 | 1 |
| Vegetables – not potatoes (tinned/frozen/ fresh) | 10 | 12 | 13 | 24 | 24 | 10 | 3 | 1 | 1 |
| Chips/fried potatoes | 17 | 14 | 21 | 29 | 9 | 4 | 3 | 1 | 1 |
| Beans or pulses (baked beans, chickpeas, dahl) | 10 | 17 | 28 | 26 | 11 | 6 | 2 | 1 | 0 |
| Fibre rich breakfast cereal, like Weetabix, Fruit ‘n’ fibre, porridge, muesli | 17 | 17 | 12 | 22 | 13 | 11 | 3 | 4 | 0 |
| Wholemeal bread or chapatis | 20 | 18 | 11 | 18 | 12 | 18 | 0 | 3 | 0 |
| Cheese/yogurt | 7 | 16 | 12 | 36 | 14 | 8 | 3 | 2 | 2 |
| Crisps/savoury snacks | 19 | 17 | 13 | 22 | 16 | 8 | 2 | 1 | 2 |
| Sweet biscuits, cakes, chocolates, sweets | 12 | 17 | 16 | 30 | 11 | 9 | 2 | 2 | 1 |
| Ice cream/cream | 42 | 30 | 14 | 3 | 3 | 6 | 0 | 0 | 1 |
| Non-alcoholic fizzy drinks/pop (not sugar free or diet) | 46 | 16 | 8 | 13 | 8 | 6 | 3 | 1 | 0 |
| Beef, lamb, pork, ham – steaks, roasts, mince or chops | 19 | 17 | 22 | 29 | 9 | 3 | 0 | 0 | 1 |
| Chicken or turkey – steaks, roasts, joints, mince or portions (not in batter or breadcrumbs) | 12 | 17 | 19 | 39 | 10 | 3 | 0 | 0 | 0 |
| Sausages, bacon, corned beef, meat pies/pasties, burgers | 22 | 16 | 27 | 30 | 2 | 2 | 1 | 0 | 0 |
| Chicken/turkey nuggets, burgers, pies, or in batter or breadcrumbs | 30 | 11 | 29 | 21 | 3 | 4 | 0 | 0 | 1 |
| White fish in batter or breadcrumbs | 41 | 24 | 21 | 8 | 0 | 3 | 1 | 0 | 1 |
| White fish not in batter or breadcrumbs | 58 | 21 | 14 | 3 | 0 | 3 | 0 | 0 | 0 |
| Oily fish (not tinned tuna) | 48 | 21 | 17 | 11 | 0 | 3 | 0 | 0 | 0 |

Supplementary Table 3: Responses to food frequency questionnaire items (in percentages) in the study sample with follow-up (n=52)

|  | Rarely/Never | | < once a week | | Once a week | | 2-3 times a week | | 4-6 times a week | | 1-2 times a day | | 3-4 times a day | | 5+ times a day | | No response | |
| --- | --- | --- | --- | --- | --- | --- | --- | --- | --- | --- | --- | --- | --- | --- | --- | --- | --- | --- |
|  | Baseline | Follow-up | Baseline | Follow-up | Baseline | Follow-up | Baseline | Follow-up | Baseline | Follow-up | Baseline | Follow-up | Baseline | Follow-up | Baseline | Follow-up | Baseline | Follow-up |
| Fruit (tinned/fresh) | 29 | 13 | 6 | 13 | 6 | 12 | 21 | 15 | 8 | 12 | 17 | 15 | 8 | 8 | 6 | 12 | 0 | 0 |
| Fruit juice (not cordial or squash) | 42 | 33 | 21 | 25 | 15 | 8 | 8 | 15 | 6 | 8 | 6 | 4 | 0 | 4 | 0 | 0 | 2 | 4 |
| Salad (not garnish added to sandwiches) | 27 | 13 | 17 | 17 | 13 | 13 | 21 | 33 | 10 | 12 | 12 | 4 | 0 | 6 | 0 | 0 | 0 | 2 |
| Vegetables – not potatoes (tinned/frozen/ fresh) | 12 | 2 | 8 | 4 | 13 | 8 | 23 | 37 | 29 | 29 | 10 | 15 | 4 | 4 | 0 | 0 | 2 | 2 |
| Chips/fried potatoes | 17 | 19 | 13 | 19 | 19 | 23 | 27 | 23 | 12 | 6 | 8 | 6 | 2 | 4 | 2 | 0 | 0 | 0 |
| Beans or pulses (baked beans, chickpeas, dahl) | 15 | 12 | 21 | 17 | 27 | 19 | 19 | 38 | 10 | 4 | 6 | 2 | 0 | 2 | 2 | 2 | 0 | 4 |
| Fibre rich breakfast cereal, like Weetabix, Fruit ‘n’ fibre, porridge, muesli | 21 | 13 | 15 | 10 | 13 | 15 | 21 | 37 | 13 | 10 | 10 | 8 | 2 | 2 | 4 | 6 | 0 | 0 |
| Wholemeal bread or chapatis | 23 | 15 | 13 | 13 | 13 | 8 | 13 | 25 | 12 | 21 | 21 | 8 | 0 | 2 | 4 | 8 | 0 | 0 |
| Cheese/yogurt | 6 | 6 | 15 | 12 | 15 | 15 | 38 | 33 | 12 | 21 | 6 | 8 | 2 | 2 | 4 | 4 | 2 | 0 |
| Crisps/savoury snacks | 19 | 21 | 17 | 25 | 17 | 17 | 19 | 21 | 10 | 6 | 12 | 8 | 0 | 2 | 2 | 0 | 4 | 0 |
| Sweet biscuits, cakes, chocolates, sweets | 12 | 10 | 15 | 8 | 19 | 27 | 29 | 25 | 8 | 13 | 10 | 12 | 2 | 6 | 4 | 0 | 2 | 0 |
| Ice cream/cream | 44 | 46 | 27 | 21 | 12 | 17 | 4 | 4 | 4 | 6 | 8 | 2 | 0 | 4 | 0 | 0 | 2 | 0 |
| Non-alcoholic fizzy drinks/pop (not sugar free or diet) | 46 | 54 | 17 | 10 | 8 | 6 | 10 | 13 | 10 | 8 | 8 | 6 | 2 | 4 | 0 | 0 | 0 | 0 |
| Beef, lamb, pork, ham – steaks, roasts, mince or chops | 19 | 12 | 12 | 8 | 21 | 17 | 35 | 48 | 8 | 8 | 6 | 2 | 0 | 4 | 0 | 0 | 0 | 2 |
| Chicken or turkey – steaks, roasts, joints, mince or portions (not in batter or breadcrumbs) | 12 | 12 | 15 | 10 | 15 | 15 | 38 | 48 | 13 | 8 | 6 | 2 | 0 | 0 | 0 | 6 | 0 | 0 |
| Sausages, bacon, corned beef, meat pies/pasties, burgers | 25 | 21 | 13 | 12 | 23 | 29 | 31 | 29 | 4 | 6 | 4 | 0 | 0 | 4 | 0 | 0 | 0 | 0 |
| Chicken/turkey nuggets, burgers, pies, or in batter or breadcrumbs | 35 | 29 | 8 | 23 | 27 | 23 | 19 | 17 | 4 | 2 | 6 | 0 | 0 | 6 | 0 | 0 | 2 | 0 |
| White fish in batter or breadcrumbs | 46 | 37 | 21 | 23 | 23 | 25 | 4 | 10 | 0 | 0 | 6 | 0 | 0 | 2 | 0 | 4 | 0 | 0 |
| White fish not in batter or breadcrumbs | 62 | 50 | 23 | 27 | 4 | 10 | 6 | 8 | 0 | 0 | 6 | 0 | 0 | 2 | 0 | 4 | 0 | 0 |
| Oily fish (not tinned tuna) | 46 | 44 | 23 | 23 | 15 | 17 | 10 | 10 | 0 | 0 | 6 | 0 | 0 | 6 | 0 | 0 | 0 | 0 |

Supplementary Table 4: Responses to food frequency questionnaire items (in percentages) in the study sample with follow-up (n=52), by concurrency of follow-up (non-concurrent follow-up at 3 months (n=14) and concurrent follow-up (n=38))

|  | Rarely/Never | | < once a week | | Once a week | | 2-3 times a week | | 4-6 times a week | | 1-2 times a day | | 3-4 times a day | | 5+ times a day | | No response | |
| --- | --- | --- | --- | --- | --- | --- | --- | --- | --- | --- | --- | --- | --- | --- | --- | --- | --- | --- |
|  | Non-concurrent follow-up | Concurrent follow-up | Non-concurrent follow-up | Concurrent follow-up | Non-concurrent follow-up | Concurrent follow-up | Non-concurrent follow-up | Concurrent follow-up | Non-concurrent follow-up | Concurrent follow-up | Non-concurrent follow-up | Concurrent follow-up | Non-concurrent follow-up | Concurrent follow-up | Non-concurrent follow-up | Concurrent follow-up | Non-concurrent follow-up | Concurrent follow-up |
| Fruit (tinned/fresh) | 36 | 5 | 0 | 18 | 21 | 8 | 14 | 16 | 14 | 11 | 14 | 16 | 0 | 11 | 0 | 16 | 0 | 0 |
| Fruit juice (not cordial or squash) | 46 | 30 | 23 | 27 | 8 | 8 | 15 | 16 | 8 | 8 | 0 | 5 | 0 | 5 | 0 | 0 | 8 | 3 |
| Salad (not garnish added to sandwiches) | 14 | 14 | 36 | 11 | 14 | 14 | 14 | 41 | 14 | 11 | 0 | 5 | 7 | 5 | 0 | 0 | 0 | 3 |
| Vegetables – not potatoes (tinned/frozen/ fresh) | 0 | 3 | 8 | 3 | 8 | 8 | 62 | 29 | 15 | 34 | 8 | 18 | 0 | 5 | 0 | 0 | 8 | 0 |
| Chips/fried potatoes | 0 | 26 | 29 | 16 | 29 | 21 | 29 | 21 | 7 | 5 | 7 | 5 | 0 | 5 | 0 | 0 | 0 | 0 |
| Beans or pulses (baked beans, chickpeas, dahl) | 7 | 14 | 14 | 19 | 36 | 14 | 43 | 39 | 0 | 6 | 0 | 3 | 0 | 3 | 0 | 3 | 0 | 6 |
| Fibre rich breakfast cereal, like Weetabix, Fruit ‘n’ fibre, porridge, muesli | 7 | 16 | 14 | 8 | 21 | 13 | 50 | 32 | 7 | 11 | 0 | 11 | 0 | 3 | 0 | 8 | 0 | 0 |
| Wholemeal bread or chapatis | 14 | 16 | 21 | 11 | 0 | 11 | 36 | 21 | 29 | 18 | 0 | 11 | 0 | 3 | 0 | 11 | 0 | 0 |
| Cheese/yogurt | 7 | 5 | 7 | 13 | 0 | 21 | 57 | 24 | 29 | 18 | 0 | 11 | 0 | 3 | 0 | 5 | 0 | 0 |
| Crisps/savoury snacks | 14 | 24 | 29 | 24 | 29 | 13 | 21 | 21 | 7 | 5 | 0 | 11 | 0 | 3 | 0 | 0 | 0 | 0 |
| Sweet biscuits, cakes, chocolates, sweets | 7 | 11 | 14 | 5 | 29 | 26 | 14 | 29 | 29 | 8 | 0 | 16 | 7 | 5 | 0 | 0 | 0 | 0 |
| Ice cream/cream | 71 | 37 | 14 | 24 | 7 | 21 | 7 | 3 | 0 | 8 | 0 | 3 | 0 | 5 | 0 | 0 | 0 | 0 |
| Non-alcoholic fizzy drinks/pop (not sugar free or diet) | 64 | 50 | 7 | 11 | 7 | 5 | 14 | 13 | 7 | 8 | 0 | 8 | 0 | 5 | 0 | 0 | 0 | 0 |
| Beef, lamb, pork, ham – steaks, roasts, mince or chops | 23 | 8 | 8 | 8 | 15 | 18 | 46 | 50 | 8 | 8 | 0 | 3 | 0 | 5 | 0 | 0 | 8 | 0 |
| Chicken or turkey – steaks, roasts, joints, mince or portions (not in batter or breadcrumbs) | 14 | 11 | 7 | 11 | 14 | 16 | 57 | 45 | 7 | 8 | 0 | 3 | 0 | 0 | 0 | 8 | 0 | 0 |
| Sausages, bacon, corned beef, meat pies/pasties, burgers | 7 | 26 | 21 | 8 | 21 | 32 | 36 | 26 | 14 | 3 | 0 | 0 | 0 | 5 | 0 | 0 | 0 | 0 |
| Chicken/turkey nuggets, burgers, pies, or in batter or breadcrumbs | 21 | 32 | 29 | 21 | 21 | 24 | 29 | 13 | 0 | 3 | 0 | 0 | 0 | 8 | 0 | 0 | 0 | 0 |
| White fish in batter or breadcrumbs | 29 | 39 | 29 | 21 | 36 | 21 | 7 | 11 | 0 | 0 | 0 | 0 | 0 | 3 | 0 | 5 | 0 | 0 |
| White fish not in batter or breadcrumbs | 64 | 45 | 21 | 29 | 14 | 8 | 0 | 11 | 0 | 0 | 0 | 0 | 0 | 3 | 0 | 5 | 0 | 0 |
| Oily fish (not tinned tuna) | 43 | 45 | 36 | 18 | 21 | 16 | 0 | 13 | 0 | 0 | 0 | 0 | 0 | 8 | 0 | 0 | 0 | 0 |
